# An Observational Study of Pulse Oximeter Alarms in a Newborn Unit at a Tertiary Facility in Kenya

**DOI:** 10.1101/2025.08.05.25332250

**Authors:** Bazil M. Masabo, Augustine W. Waswa, Jesse Coleman, Morris Ogero, Grace Irimu, Amy Sarah Ginsburg, Dorothy Chomba, Millicent Parsimei, Cynthia Shitote, Ferdinand Okwaro, June K. Madete, William M Macharia, J Mark Ansermino

**Affiliations:** Kenyatta University, London, UK; University of British Columbia, London, UK; Agha Khan University Nairobi, London, UK; University of Nairobi, London, UK; University of Washington, London, UK; London School of Hygiene and Tropical Medicine, London, UK

## Abstract

**Background:** Continuous pulse oximetry monitoring of neonates in the intensive care unit improves clinical outcomes. However, monitoring will only be effective if actions are taken based on the results of appropriate monitoring. While threshold alarms are used to alert providers about clinical deterioration, alarm fatigue significantly degrades the effectiveness of alarms if the thresholds are not adapted based on the clinical status of each neonate, including changes seen in neonatal physiology.

**Methods:** As part of a quality improvement project aimed at reducing the risk of apnea of prematurity, which included caffeine citrate administration, we conducted an observational study of alarms during the implementation of continuous pulse oximetry in a newborn unit (NBU) of a tertiary teaching and referral healthcare facility in Kenya. Default alarm thresholds were set at 85 to 96% for oxygen saturation (SpO_2_) and 90 to 200 beats per minute (bpm) for pulse rate (PR).

**Results:** Among 49 neonates with a median birth weight of 1.3 kilograms (IQR,329 grams) at admission, 16,212 hours of data were recorded. Respiratory support was provided on 28% of patient days. The median number of alarms per hour per neonate was 12 (IQR,40) visual and 9 (IQR,21) audible alarms. Half of the SpO_2_ values lay outside the thresholds, with 44% exceeding the upper threshold of 96%, with a median duration of 20 (IQR,72) seconds. The mean for HR was 1 (SD,1.17) alarm per hour per neonate, with 2% of the data lying outside the thresholds.

**Conclusions:** The thresholds for SpO_2_ resulted in a high alarm burden. A short alarm delay could have significantly reduced this burden. Setting an upper threshold of 96% in clinically stable babies resulted in a large volume of unnecessary alarms. Adjusting thresholds based on individual neonatal characteristics, including consideration of the need for respiratory support, is recommended. Effective monitoring of neonates requires individualized alarm thresholds and additional smart algorithms specifically designed to detect clinical deterioration.

## Background

Preterm birth, defined as live birth before 37 weeks of gestation, is among the leading causes of neonatal mortality and is associated with long-term physical, neurodevelopmental, and socioeconomic effects (Ohuma et al., 2023). In 2020, an estimated 13.4 million preterm births were recorded worldwide, which is more than 1 in 10 infants. Despite being a global issue, there are regional disparities in the survival rates of preterm infants. Over 90% of preterm infants born in low-income countries, where most preterm births occur, die within the first few days of life (World Health Organization, 2023). The death rate is less than 10% in high-income countries. In Kenya, the neonatal mortality rate is 21 deaths per 1,000 live births, with preterm birth the leading cause of death (KDHS 2022 Key Indicator Report.pdf).

Preterm-born children are susceptible to adaptation problems in the early neonatal period and to poor outcomes due to the immaturity of the respiratory, cardiovascular, and especially cerebrovascular systems (Zivaljevic et al., 2024). Compared to term infants, preterm infants are four times more likely to die in the first 28 days of life, with mortality rates increasing proportionally with decreasing gestational age (GA) or birth weight (Razeq, Khader, & Batieha, 2017). Extremely preterm infants frequently require some form of respiratory assistance to facilitate cardiopulmonary transition in the first hours of life (Tana et al., 2023). Respiratory distress syndrome (RDS) affects about 80% of neonates born at 28 weeks GA, and this percentage increases to 90% at 24 weeks GA.

The vulnerability of preterm infants typically necessitates critical, individualized, and continuous care in a neonatal intensive care unit (NICU) that can provide access to appropriate medical technologies. NICU medical technologies can enable lifesaving care through continuous monitoring, pulse rate (PR), oxygen saturation (SpO_2_), electrocardiography, and brain activity (Taha, Simpson, & Sharkey, 2023).

The high number of physiological parameters requiring monitoring in the NICU and an expanding array of medical technologies have caused an increased frequency and volume of medical device alarms. Research shows that alarm fatigue, defined as a condition of sensory overload for healthcare providers (HCPs) exposed to an excessive number of alarms (Blake, 2014), is a serious problem in NICUs that can impact the quality of care and patient outcomes (Sendelbach & Funk, 2013). Alarm fatigue can contribute to negative perceptions among HCPs, which can impede the delivery and quality of care and lead to an increased risk for adverse patient outcomes (Ruppel et al., 2018; Patel et al., 2022). Given that over 70% of clinical alarms do not require clinical intervention (Sendelbach & Funk, 2013), overburdened clinicians can become desensitized to medical device alarms and subsequently less responsive to clinically significant alarms.

There is growing interest in understanding and appreciating the importance of addressing alarm hygiene and how it contributes to alarm fatigue (Ruskin & Hueske-Kraus, 2015). Although medical device alarms are crucial for patient safety and effective clinical care, minimal evidence-based strategies exist to improve alarm hygiene. Different approaches have been tried, including an educational package on alarm management (the number of alarms, response to alarms, and appropriateness of settings) that resulted in a significant decrease in the number of alarms and an improvement in the number of times where appropriate alarm settings were used (Hedda et al., 2021). Other interventions include the adjustments of alarm thresholds and delay settings according to the patient’s condition (Welch, 2011). An evidence-based approach that balances patient safety with alarm sensitivity and specificity is critical for all interventions.

In an effort to provide evidence to improve alarm hygiene techniques for more effective management of neonates receiving continuous monitoring, we conducted a secondary analysis of alarm data obtained from pulse oximeters used in monitoring preterm neonates in a clinical feasibility study focused on using caffeine citrate to manage apnea of prematurity (AOP) in a single facility tertiary-care newborn unit (NBU) in Nairobi, Kenya (Irimu et al., 2023).

## Methods

### Study Design

As part of a larger quality improvement study that included administration of caffeine citrate and continuous monitoring for neonates at risk of AOP, this observational study of pulse oximeter alarms included a 4-month formative research phase followed by the development of an AOP prototype clinical-care bundle. Implementation and improvement of the clinical-care-bundle prototype were done in the second phase. The baseline pulse oximetry data was used to provide contextualized insights on care practices within the NBU to inform the development of a context-sensitive AOP prototype clinical-care bundle.

Recognized as a reference site for newborn care standards in Kenya, this NBU in a tertiary teaching and referral healthcare facility in Nairobi, Kenya, has considerable potential to influence clinical practices broadly. The NBU staff included five neonatologists, seven neonatology fellows, one pediatrician, three medical officers, and 12-15 pediatric residents. One medical officer and one resident did night coverage. The NBU had a complement of 80 nurses working in shifts, with an average of 13 on each shift. Training sessions on the management of AOP were conducted for all staff, including nurses, residents, medical officers, consultants, and nutritionists.

Typically, 150 neonates are admitted monthly to the NBU, which has an average bed capacity of 60 neonates, with neonates often sharing essential equipment such as cots, radiant warmers, and incubators. Annually, 13% of NBU admissions are very low birth weight neonates (1000-1499 grams), with 85% admitted on the day of birth (Source: Hospital NBU Database). For this study, a convenience sampling of consecutive neonates admitted to the NBU during the defined study period who were born less than 34 weeks gestational age (GA), or with a birth weight less than 1500 grams where the GA was unknown, provided the estimated GA was less than 34 weeks as determined using New Ballard scoring, was screened for enrollment. Neonates receiving caffeine citrate or aminophylline for AOP were enrolled if their caregivers provided written informed consent. Routine continuous pulse oximetry was implemented during the study, and data from the devices were captured and analyzed.

### Study Procedures

Rad-G™ pulse oximeters with reusable Masimo Yi sensors were used for continuous monitoring. Formal training in using the pulse oximeters was provided to all nursing staff. The devices were connected to the neonates by the NBU nurses as soon as the neonates were admitted to the NBU. Routine clinical hygiene measures were followed, including strict hand hygiene during attachment and detachment of the sensors and cleaning the pulse oximeters and sensors when transferring from one baby to another. The study protocol recommended that the location on the neonate’s body where the pulse oximeter sensor was attached should be changed every 3 hours, but this was not consistently implemented.

Based on clinical consensus, default alarm thresholds were set during device setup (Table 1). These settings could be changed if the neonate was not receiving oxygen or for any other clinical indication. The study protocol recommended that the upper threshold be disabled for neonates not on oxygen, but this was not consistently followed.

**Table 1.** Default alarm thresholds.

| Parameter | Threshold |
| --- | --- |
| Sensitivity | Normal |
| Averaging time | 8 seconds |
| Rapid desaturation | 10% |
| Alarm delay | 15 seconds |
| Fast Sat | OFF |
| SpO<sub>2</sub> | Upper threshold: 96%<br>Lower threshold: 85% |
| PR | Upper threshold: 200 bpm<br>Lower threshold: 90 bpm |

Based on the device identifiers, the study nurses recorded which neonate was attached to which device each day. At the end of each day, data were transferred from each device to a universal serial bus (USB) drive and then to a laptop computer running the Masimo Trace Software^@^. Data at 0.5 hertz (Hz) resolution was exported into comma-separated values (CSV) files for further analysis.

## Data Analyses

R Studio 4.2.3 and dplyr, tidyverse, ggplot, gridExtra, lubridate packages were used to clean and analyze data and perform statistical analyses. Operations included merging CSV files by patient identification number (PID), removing irrelevant columns, and refining the dataset by eliminating empty cells. We replaced missing values with null values. Clinical events were split into six columns for each unique alarm entry category for detailed analysis. The description of relevant terms is captured below. (Table 2).

**Table 2.** Description of device terminologies.

| **Term** | **Description** |
| --- | --- |
| Pulse oximeter alarm | An alert triggered by the pulse oximeter when a measured value falls outside a set range |
| Audible alarm | An audible alert sounded by the pulse oximeter |
| Visual alarm | A visual alert displayed by the pulse oximeter |
| System alarm | An audible and/or visual alarm due to a system event, such as a sensor being detached or motion detected |
| Clinical alarm | An audible and/or visual alarm due to any out-of-range parameter |
| SpO<sub>2</sub> | Percentage of oxygenated blood measured with pulse oximetry |
| Pulse rate (PR) | Number of heartbeats per minute measured with pulse oximetry |
| Perfusion index (PI) | Pulse oximetry-specific ratio of pulsatile volume of blood flow compared to a baseline measurement; used as a surrogate for peripheral perfusion to guide appropriate sensor placement |

We excluded rows of data with errors occurring when continuous pulse oximetry was not available. We focused our analysis on SpO_2_ and PR, utilizing R’s quantitative tools and visualizations (histograms, bar graphs, pie charts). In addition, we removed invalid entries for SpO_2_ (“Invalid functional SpO_2_”) and PR (“Invalid PR”), together with alarms due to low SpO_2_ signal identification and quality indicator (SIQ).

We counted visual and audible alarms by considering the occurrence of a visual, audible, or audio-visual alarm. Each cell of the dataset represented two seconds of monitoring. We calculated alarm duration by counting consecutive occurrences of unique events and multiplying by two seconds. A consecutive occurrence of a unique event represented one alarm unless interrupted by the occurrence of a different event or a gap of more than 30 seconds between occurrences of the same event.

## Results

Of the 49 neonates enrolled in the study, there were more males (31; 63%) than females (18; 37%), and the median birth weight at admission was 1.3 kilograms (IQR,329 grams). Seven (14%) neonates died during the study. Respiratory support was provided during 28% of patient days, including intubation and mechanical ventilation during 194 (13%) days and continuous positive airway pressure (CPAP) during 219 (15%) days. We recorded 16,212 hours of monitoring data overall. Continuous monitoring durations of SpO_2_ and PR varied across neonates from a minimum of 16 hours up to a maximum of 41 days, with a median monitoring duration of 11.2 (IQR,14.2) days. We excluded 9.97% of the total duration of monitoring data for poor data quality, mainly due to system events, including pulse searching, sensor detachment, low perfusion index, and cable disconnection. These system events frequently resulted in non-values (74.8% for SpO_2_ and 73.6% for PR). The poor-quality data contained 31,058 (5.4%) alarms in total.

### Overall alarm burden

There were 565,413 clinical alarms during monitoring, with 87% (493,577) lasting 30 seconds or less. Audible alarms accounted for 44% of the total alarm burden and had a median duration of 8 (IQR,36) seconds. The alarms over 30 seconds lasted a median duration of 78 (IQR,124) seconds. Audible alarms lasted longer than visual alarms, and almost all alarms that lasted more than two minutes were audible. Alarms of durations more than five minutes and 10 minutes accounted for 4.2% and 2.1% of audible alarms, respectively.

Each neonate experienced a median of 9,993 (IQR 15,600) alarms throughout monitoring, translating to a median duration of 97 (IQR,123) alarm hours. The highest hourly count was 201 alarms, and the most alarm-burdened day had 2,179 alarms. The median alarm density (number of alarms per hour) per neonate was higher for visual (12 (IQR,40)) than for audible (9 (IQR, 21)) alarms. (Figure 1).

**Figure 1.**
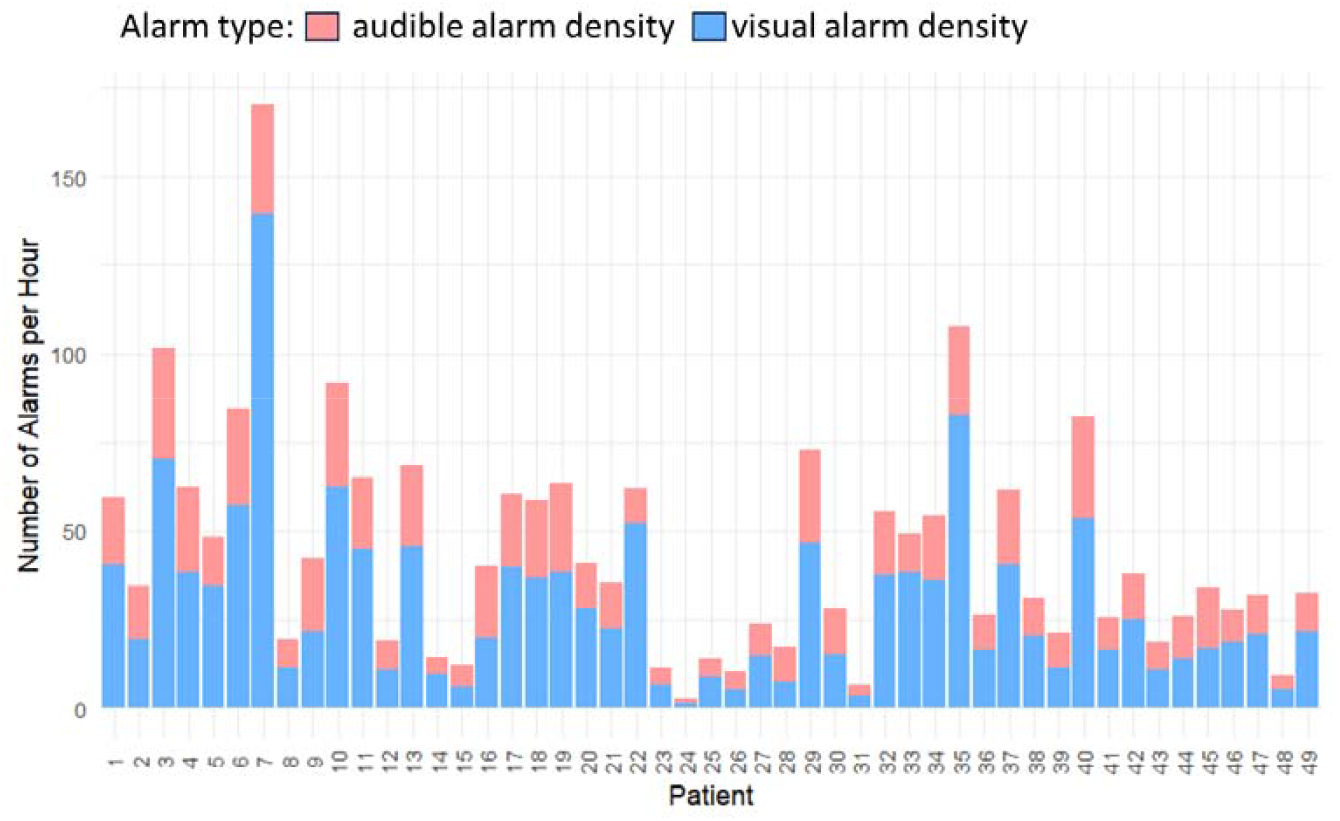
Per patient alarm density

### Oxygen saturation alarms

With an overall median SpO_2_ value of 96% (IQR,5%), we recorded 98,047 SpO_2_ alarms, 56,312 (57.4%) high, and 41,735 (42.6%) low alarms. Of the 50% recorded SpO_2_ values that were outside the set threshold range, 44.5% were greater than the upper threshold (36% with 100% SpO_2_; 20% with 99% SpO_2_; 22% with 98% SpO_2_; and 22% with 96% SpO_2_). The median duration of SpO_2_ alarms was 20 (IQR,72) seconds. Most alarms - 80% of low alarms and 60% of high alarms - were less than a minute long, while longer duration (above 4 minutes) alarms were predominantly high SpO_2_ alarms (over 15%). (Figure 2). Overall, 5.78% of the SpO_2_ values were below 85%, leading to a median of 1.7 (IQR,0.6) alarms per hour per neonate. On average, each neonate had a median of 1,643 (IQR,2950) SpO_2_ alarms, with the highest number being 13,680 alarms. The median SpO_2_ alarm burden for both low and high alarms was 4 (IQR,5) alarms per hour per neonate. There was a higher number of SpO_2_ alarms than PR alarms. (Figure 3).

**Figure 2.**
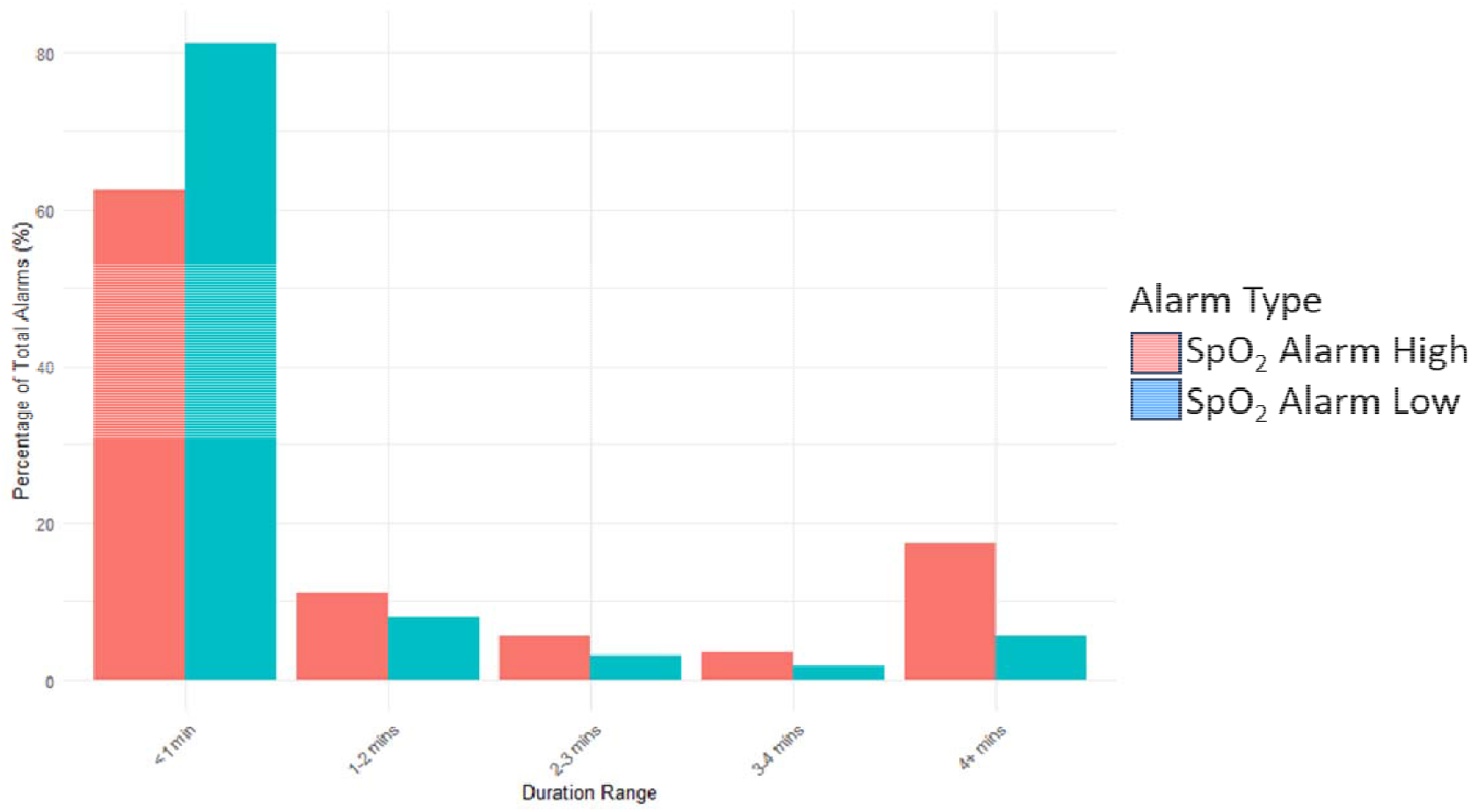
Percentage of SpO_2_ alarms with different durations as a percentage of total SpO2 alarms.

**Figure 3.**
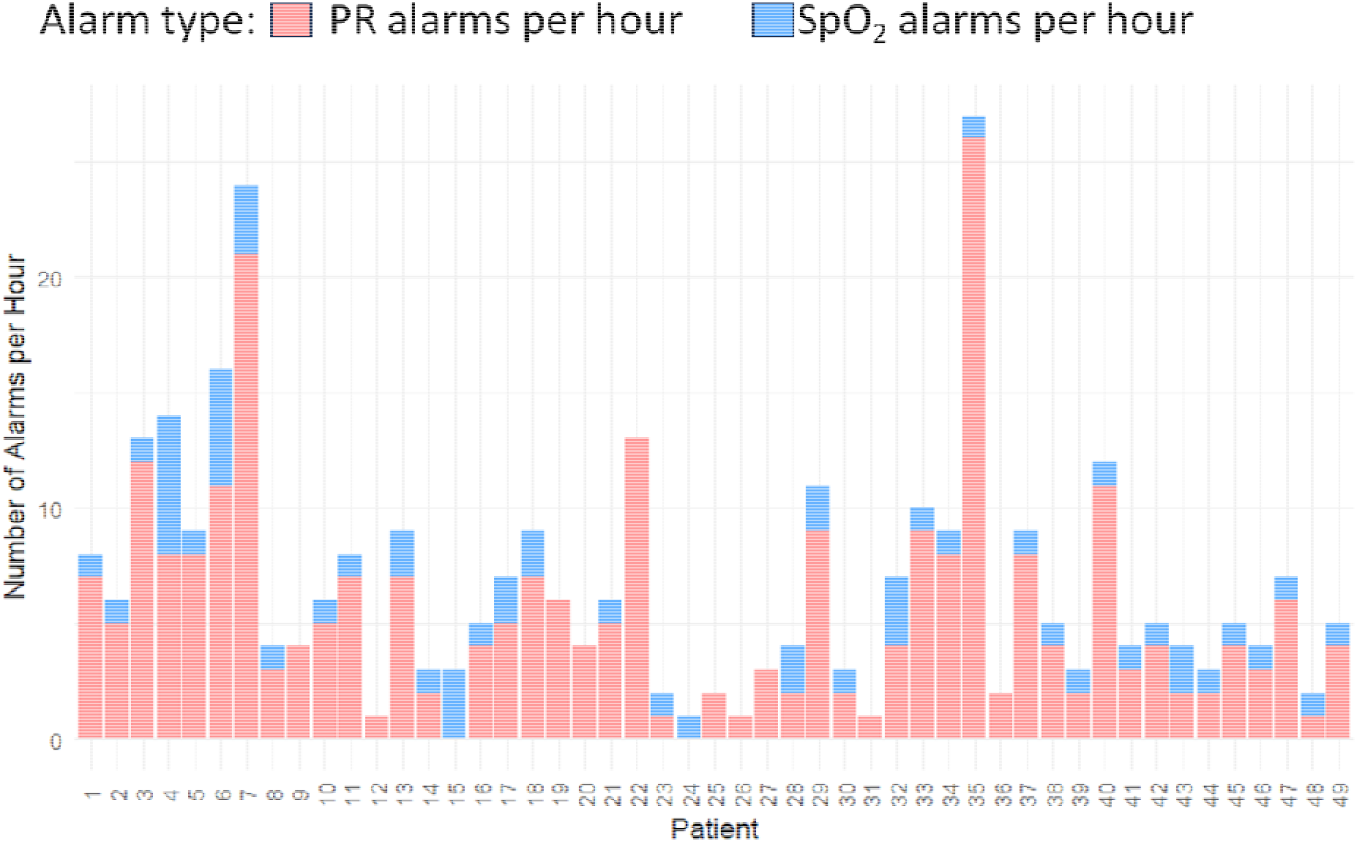
Per patient alarm densities

### Pulse rate alarms

With a mean (standard deviation (SD)) PR value of 158 (19.87) beats per minute (bpm) throughout monitoring, we recorded 18,218 PR alarms, 58.5% high PR and 41.5% low PR alarms. The mean (SD) alarm density was 1 (1.17) PR alarm per hour per neonate, with 2% of the data lying outside the thresholds. The median number of alarms per neonate was 199 (IQR,347), and each neonate experienced, on average, 371 PR alarms throughout monitoring with a median of 71 (IQR,156) alarms per day per neonate. PR alarms contributed a smaller portion of the total alarm burden than SpO_2_ alarms. (Figure 4). When monitoring PR, 14,096 alarms (77%) lasted shorter than a minute. High alarms were the majority, both for alarms with durations below a minute and for longer-duration alarms. The median duration for PR alarms was 18 (IQR,44) seconds.

**Figure 4.**
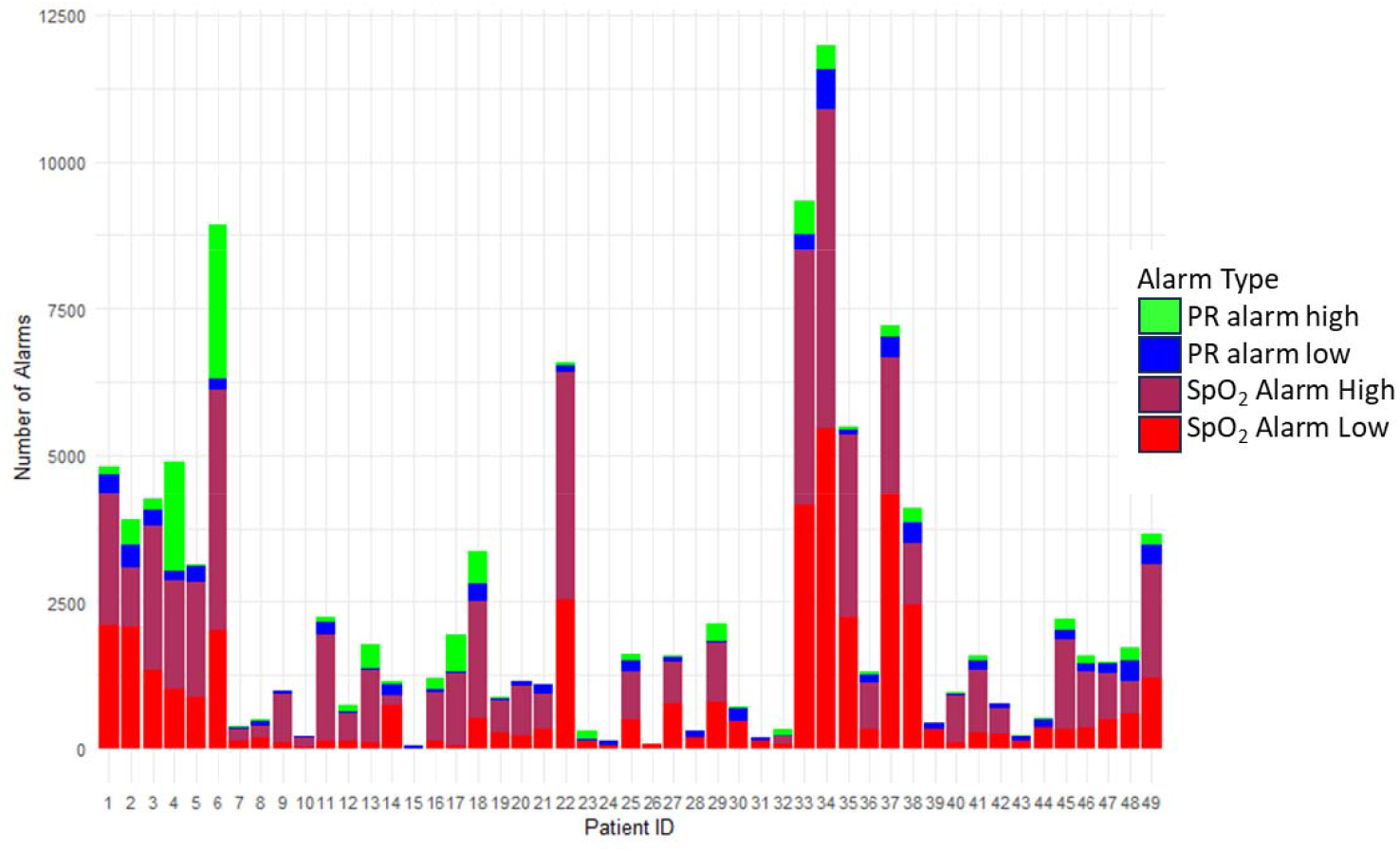
Number of alarms per patient by alarm type

## Discussion

The introduction of continuous pulse oximetry into the NBU resulted in a significant alarm burden, primarily of short-duration alarms. Over 85% of alarms lasted one minute or less, with many lasting only two seconds. Although most were visual alarms, 43% were audible alarms. Multiple small changes in SpO_2_ and PR were responsible for many of the alarms, and not all were likely indicative of clinically significant events. Since neonates are prone to high alarm burdens, transient changes in SpO_2_ and PR could be due to intrinsic neonatal physiological fluctuations, patient movements, sensor interference, and poor signal quality. Many alarms were self-resolving without clinical intervention, thereby contributing to a large proportion of non-actionable alarms.

Frequent non-critical alarms that are clinically irrelevant can create habituation among HCPs, leading to insensitivity to alarms and reduced responsiveness to real clinical events as well as increased perceived workload (Staff, 2022). Persistent exposure to non-actionable alarms can result in slower response times and missed or delayed interventions during emergencies, with serious and sometimes deadly consequences (Sanz-Segura et al., 2019; Lu, 2024).

Furthermore, the frequent arousal of neonates may have deleterious effects on infant growth, development, and health. Sleep is critical for physical growth, brain maturation, and autonomic regulation in neonates. Frequent awakenings due to alarms have been linked to increased cortisol levels, indicating heightened stress, which can interfere with immune function and neurodevelopmental progress. Frequent and unnecessary disruption of sleep cycles caused by audible alarms may have long-term developmental implications for neonates (Patel et al., 2022).

### Alarm hygiene

#### Introducing alarm hygiene

Minimizing non-actionable alarms is critical to appropriate alarm hygiene. Alarm hygiene is a set of strategies and practices implemented to manage and reduce the frequency of false alarms in systems, particularly in industrial and healthcare settings (OSHA, 1998). Effective alarm hygiene ensures that alarms are meaningful and actionable and improve safety and efficiency. There is an intricate balance between sensitivity and specificity to achieve a high signal-to-noise ratio and avoid missed events without unnecessary alarms.

#### Increasing alarm delay

With 87% (493,577) of alarms in our study lasting 30 seconds or less, increasing the alarm delay is one effective method to reduce the alarm burden. A delay in the time taken for an alarm to trigger allows transient changes in the patient parameter to self-correct. This can filter out non-actionable alarms due to brief fluctuations in the measured parameter or interferences from patient movements. An alarm delay of 15 seconds can reduce alarm frequency by 70% (Welch, 2011). Alarm delays allow improvement in alarm specificity without affecting a reasonable clinical response time (Welch et al., 2016). Moreover, modifications of alarm delays to suit patient conditions could further reduce the overall number of alarms in clinical environments (Jämsä, Uutela, Tapper, & Lehtonen, 2021), particularly for high-frequency alarms like low SpO_2_ and SpO_2_ probe off.

We observed in our study that of the alarms lasting less than 30 seconds, the greatest proportion lasted less than 10 seconds. This indicates that many alarms may not be clinically significant but could still contribute to alarm fatigue. Therefore, implementing a 10-second time delay as a filter could mitigate the issue by reducing transient, momentary alarms that often do not reflect critical patient events.

#### Modifying oxygen saturation alarm thresholds

Modifying oxygen saturation alarm thresholds according to patient conditions and treatments presents another technique for reducing the frequency of non-actionable alarms. A 2% change in the lower SpO_2_ threshold could reduce the alarm burden by up to 50% (Jämsä, Uutela, Tapper, & Lehtonen, 2021). The results suggest that optimizing SpO_2_ thresholds may further improve the specificity of alarms, minimize alarm fatigue, and enhance overall patient monitoring efficiency (McCauley et al., 2021).

The SpO_2_ alarm thresholds used in this study captured a substantial proportion of data outside the thresholds, with the overwhelming majority exceeding 95%, the upper threshold, and 38% due to SpO_2_ values of 100%. For premature neonates on supplemental oxygen, a high SpO_2_ value indicates excessive oxygen administration, which can increase the risk of retinopathy; however, for neonates on room air, a high SpO_2_ value would be clinically reassuring. Thus, we recommend a default upper alarm threshold of 95% (Askie et al., 2018) for those on supplemental oxygen, while we would recommend no upper threshold for those not on oxygen.

Reducing the low SpO_2_ alarm threshold from 90% to 85% can decrease alarms by 75% (Welch, 2011). Less than 6% of alarms in our study were for SpO_2_ values below 85%, the lower threshold. Adjusting the lower threshold to 80% could significantly reduce the proportion of excluded data. A theorized minor adjustment (e.g., 80.2%) had a pronounced impact, excluding only 3.4% of the data compared to 6% at 85%. This suggests a high sensitivity of data distribution around the lower threshold. Clinically, this indicates the importance of selecting alarm thresholds that balance sensitivity (capturing relevant hypoxemia events) and specificity (avoiding unnecessary alarms that may not represent critical events). We recommend a lower threshold of 80% (Askie et al., 2018), but with individualized threshold adjustment for infants with SpO_2_ values consistently below this threshold despite context-appropriate interventions.

#### Modifying pulse rate alarm thresholds

Complementary to the low SpO_2_ alarm, the low PR alarm reflects the neonate’s physiological response to hypoxemia and could be used to escalate the urgency of the SpO_2_ alarm. A significant drop in SpO_2_ initiates a series of compensatory mechanisms to ensure adequate organ oxygenation. Increased cardiac output is initially common, but in severe cases, a bradycardic response takes over as the body’s final attempt to reduce oxygen consumption. In our study, the PR thresholds of 90 to 200 bpm resulted in 2% of the data being outside the thresholds. For the low PR threshold of 90 bpm, only 0.92% of the data fell below the threshold compared to 1.1% at a theoretical threshold of 98%. We recommend a low PR threshold of 90 bpm.

#### Combination of both methods

A combination of both optimizing alarm thresholds and utilizing alarm delays provides a higher reduction in the alarm burden, which can be critical in balancing the competing priorities of patient safety and alarm fatigue mitigation. Lowering alarm limits to 88% with a 15-second delay can reduce alarms by over 85% (Welch, 2011). These two settings offer significant alarm reduction while preserving actionable alarms and thus maintaining patient safety, which is a focus of alarm hygiene.

#### Argument for data-driven adjustments

These data-driven interventions could lead to daily alarm densities of less than 10 alarms per day even in busy settings (Berg et al., 2023). This reduction highlights how modest adjustments to alarm settings can meaningfully impact alarm frequency in busy clinical environments. Furthermore, analyses of several alarm burden reduction methodologies indicate that critical monitoring, such as during events leading to code blue or high-acuity transfers, is not compromised (Berg et al., 2023) by data-driven adjustments to alarm delays to suit patient conditions.

#### Argument for organizational change

Optimizing and individualizing alarm thresholds and delays can reduce the alarm burden, but also should be combined with organizational change, using traditional quality improvement methodologies, to optimize responsiveness to alarms (Sendelbach & Funk, 2013). Other strategies to reduce alarm burden include selecting appropriate technologies, such as sensors specifically designed for use with premature neonates, and optimizing signal quality by proper placement of sensors (Winters et al., 2018). Excluding external light, keeping the environment warm, preventing motion of the sensor and cable, and using algorithms to track the dynamic changes in monitoring, in addition to thresholds, are also considerations.

## Limitations

Our study was limited by the convenience sampling strategy, a small sample size, and a potential lack of generalizability to other patient populations and healthcare settings. In addition, in our analysis, we classified 9.97% of the data as poor quality, making up about 31,053 (5.4%) alarms. Common causes of poor-quality data were system events, including pulse searching, sensor detachment, low perfusion index, and cable disconnection. Alarms triggered by such events can contribute to the alarm burden. Since the study enrolled neonates who frequently moved their hands and feet, sensor detachments are expected to be common. Therefore, good practice for sensor placement is necessary.

## Conclusions

In the context of our study, these results reinforce the idea that optimizing alarm settings is an effective strategy for reducing alarm burden. Our findings also underscore the importance of data-driven adjustments, where thresholds are tailored based on clinical priorities and patient population needs. These data-driven adjustments ensure that critical events remain detectable while reducing the noise of non-actionable alarms.

Since premature neonates are prone to high alarm burdens due to their unique physiology and care needs, data-driven alarm management strategies are more critical in the NICU. Neonate-specific alarm thresholds and well-adjusted alarm delays can improve the specificity and sensitivity of monitoring devices. When device alarm settings are coupled with appropriate organizational adaptations, patient care and safety are enhanced while HCP exposure to excessive alarms is mitigated.

## Data Availability

All data produced in the present study are available upon reasonable request to the authors.

## Acknowledgements

We are grateful to the Bill & Melinda Gates Foundation for funding this project (Proposal No. [INV-035872-2021]). We are also grateful to the Kenyatta National Hospital administration and the NBU clinical teams, as well as the caregivers of the neonates. This is not forgetting all the individuals whose valuable support and contributions were instrumental in the successful completion of this study.

## Author contributions

BM: Formal analysis, Software, Visualization, Writing – original draft, Writing – review & editing. AW: Formal analysis, Software, Visualization, Writing – original draft, Writing – review & editing. JC: Resources, Software, Writing – review & editing. MO: Data Curation, Software, Writing – review & editing. ASG: Conceptualization, Project administration, Writing – review & editing. DC: Project administration, Investigation, Data Curation, Writing – review & editing. MP: Investigation, Data Curation, Writing – review & editing. CS: Investigation. JM: Formal analysis, Visualization, Writing – original draft, Writing – review & editing. FO: Conceptualization, Project administration, Methodology, Funding acquisition, Resources. GI: Conceptualization, Project administration, Methodology, Funding acquisition, Resources, Writing – review & editing. WM: Conceptualization, Project administration, Methodology, Funding acquisition, Resources, Writing – review & editing. MA: Conceptualization, Project administration, Methodology, Writing – review & editing. All authors provided feedback and review of the manuscript.

